# Bootstrapping surgeon labeling campaigns with x_0_-diffusion: label-efficient vessel segmentation of catheter-based angiograms

**DOI:** 10.64898/2026.11.13.26368001

**Authors:** Neelay Ranjan, Shantanu Dev, Andrew Alexander Gonzalez

## Abstract

**Objective:** Assess which architecture is the most label-efficient for vessel segmentation of catheter-based angiograms, and whether a diffusion model trained with extreme data scarcity could serve as a scaffold for expert annotation.

**Materials and Methods:** *x*_0_-diffusion, SAM, DeepLabV3, ResNet-UNet, ViT-DPT, and ResNet-ViT hybrid were compared at 16, 32, and 80 labeled images from the 500-image Dr-SAM dataset under 5×5-fold repeated cross-validation, using one-sided paired Wilcoxon signed-rank tests with Holm correction. A vascular surgeon’s annotation time, from scratch and when correcting *x*_0_-diffusion predictions, was also measured.

**Results:** At 16 training images, *x*_0_-diffusion achieved the highest Dice (0.882) and IoU (0.795) of the six models, outperforming every competitor in all 25 paired runs on Dice, by 0.047 over zero-shot SAM and 0.226 over DeepLabV3. The advantage narrowed as labels increased: at 32 images *x*_0_-diffusion was not significantly different from the two strongest convolutional models, and at 80 images those models scored marginally higher (0.923 and 0.920 vs. 0.910). Correcting 16-image *x*_0_-diffusion predictions on five test angiograms averaged 11.1 surgeon-minutes per image versus 20.6 from scratch.

**Discussion:** The advantage of *x*_0_-diffusion is label efficiency, not peak accuracy, with the strongest convolutional models needing twice the labels to match it. It rarely fails where zero-shot SAM fails, so at the start of an annotation campaign, correcting its predicted masks could cut surgeon annotation time by an estimated 37 to 45%.

**Conclusion:** With very few labels, *x*_0_-parameterized diffusion is the strongest vessel segmenter tested and a candidate scaffold for expert annotation.

**Layman Summary:** Segmenting vessels on an angiogram manually takes a vascular surgeon about twenty minutes per image. This study shows that an *x*_0_-diffusion model trained on only 16 angiograms draws outlines good enough to correct rather than redraw, which took the surgeon about half as long on five test images.

---

In the United States, peripheral artery disease (PAD) affects an estimated 10 to 12 million adults over the age of 40 years (and nearly 236 million people worldwide). ^1^ Catheter-based angiography (CbA) uses x-rays to visualize contrast dye that is injected into blood vessels and plays a unique role in the diagnosis and treatment of PAD. As a diagnostic study, CbA remains the gold-standard for definitive diagnosis particularly in cases of diagnostic uncertainty. CbA facilitates operative planning due to its ability to reliably resolve blood vessels below the knee and simultaneously provide both anatomic and physiologic data. These attributes make it vital in assessing target vessels for surgical bypass. Finally, CbA serves as an interventional platform for numerous intraarterial interventions including balloon angioplasty, stent placement, lithotripsy, atherectomy, “over the wire” embolectomy, and catheter directed thrombolysis. ^1–3^

## BACKGROUND AND SIGNIFICANCE

Despite high clinical value and wide availability of raw images, CbA remains a largely untapped data source and unexplored modality in machine learning for multiple reasons. First, there is significant variation in label quality and terminology. Second, images are not routinely annotated in a way that is easily digestible by machine learning pipelines (i.e. “arrow signs”). Finally, angiographic interpretation requires specialized training and is only routinely performed by three relatively small specialties (vascular surgery, interventional cardiology, interventional radiology).

Our ability to capitalize on the rich clinical information contained in angiography studies is constrained by cost, time, and scarcity of experts. ^4,5^ This “annotation bottleneck” remains the critical gap in developing meaningful image-based clinical decision support for PAD. In other domains progress is often limited by model capabilities. In contrast, for advancing AI-enhanced PAD care, we are limited by the efficiency of end-to-end annotation to inference pipelines.

With advances in computer vision, several published works address how to handle imaging based tasks where data constraints are an issue. ^6,7^ For our paper, we explore label-efficient segmentation to identify vessels within CbAs. This involves providing a limited set of annotated examples in order to learn a new classification of a structure within an image. A related setting, few-shot segmentation, which learns a new class from a small support set at inference time, has seen several advances over the past few years. ^8^ A notable one is Meta’s Segment Anything Model (SAM), a generalized segmentation model driven by input prompts such as points or bounding boxes, ^9^ which has been adapted to the medical domain. ^10^ However, it still requires manual prompts, which cannot be reused for downstream ML tasks, and it struggles to capture complex arterial branching as seen in CbAs. ^11,12^

Diffusion models have been responsible for some of the state of the art image generation tasks. ^13,14^ Instead of treating segmentation as a discriminative task, the problem is reformulated as a generative task. This method has been shown to be effective in label-efficient settings. ^15–17^

## OBJECTIVE

This paper determines which of six architectures is the most label-efficient for vessel segmentation on catheterbased angiograms, and how each behaves as labels are added. Finally, we determine whether the most label-efficient architecture can bootstrap a labeling campaign and reduce the surgeon-hour cost of starting one.

## MATERIALS AND METHODS

### Dataset

The Dr-SAM benchmark dataset ^12^ contains 500 X-ray angiograms of the infrarenal abdominal aorta and iliac arteries with the vascular masks and the bounding boxes marked by two different radiologists (Figure 1); 450 images are 386 × 448 pixels and 50 are 819 × 950. Grayscale images were triplicated across channels for the CNN and ViT encoders, and masks were provided as single-channel binary images.

**Figure 1.**
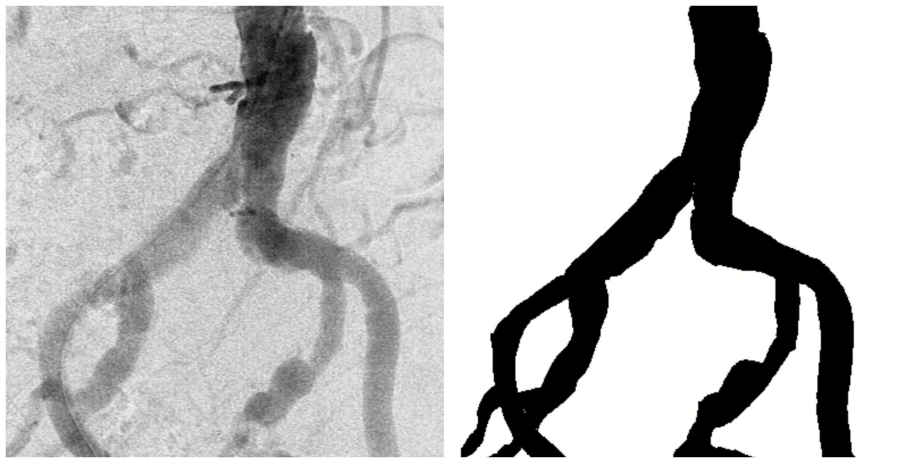
Example angiogram and expert vessel mask from the Dr-SAM dataset.

Alt text: An X-ray angiogram of the infrarenal aorta and iliac arteries on the left, with the corresponding expert-drawn binary vessel mask beside it on the right.

No data augmentation was used (e.g., rotation, contrast, or affine transformations). Image-mask pairs were resized to 384 × 384 and normalized to zero mean and unit variance to match the input conventions of the en-coders, with the size being constrained by the inflexible input size of ViT-based architectures. ^18^ No inclusion or exclusion criteria were applied, and all 500 image-mask pairs were used. The dataset is anonymized. ^12^

### Task definition

The task is 2D binary segmentation of blood vessels in angiograms, where the input is a 3 × 384 × 384 image (with positive points for SAM) and the output a 384 × 384 binary vessel mask. Evaluation criteria were Dice and IoU, ^19,20^ which are computed per image on the test dataset. For any given predicted mask *p* and ground truth *g*,

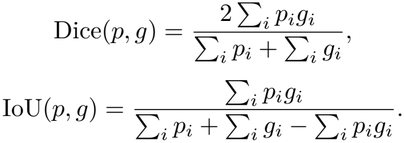

### Model architectures

The architectures were chosen to span a range of inductive biases highly relevant to label-efficient segmentation. ResNet-UNet, a convolutional model, has strong local context; at the other end, ViT-DPT, a transformer, has global context. All are compared to an iterative refinement diffusion model with the *x*_0_ parameterization as its target. Trainable parameter counts are 56.6 M for the diffusion UNet, 32.5 M for ResNet-UNet, 39.6 M for DeepLabV3, 122.1 M for ViT-DPT, 154.6 M for the ResNet-ViT hybrid, and 93.7 M for SAM ViT-B, so the diffusion model is larger than both convolutional baselines and smaller than the transformer-based ones.

### Diffusion

The diffusion model takes a 4-channel input, broken down into a 1-channel noisy mask concatenated with the 3-channel clean angiogram, which is the conditioning signal. The model architecture is based on the UNet2DModel, ^21^ which has channel sizes of (64, 128, 256, 512) as a trade-off between capacity and computecost. The noise schedule is the DDPM linear beta schedule ^22^ with 1000 training timesteps. Training involves sampling a timestep *t*, noising the ground truth mask *x*_0_ at the timestep in the forward process

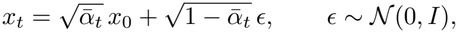

concatenating *x*_*t*_ to the image, and finally predicting the clean mask.

Inference starts from a noise mask and runs 100 DDPM steps with the image concatenated at each step; fewer steps than the 1000 training timesteps are used to reduce inference time. The final prediction is thresh-olded at 0.5 to produce the binary mask on which all metrics are computed.

As an ablation of the prediction target, an *ϵ*-diffusion reparameterization with the same architecture, schedule, conditioning, and timestep budget was also trained; because it predicts noise, it cannot use a mask-space loss and was trained with MSE in noise space.

### *x*_0_-diffusion

The model outputs clean mask logits directly (an *x*_0_ prediction) rather than the additive noise (*ϵ* prediction) of the original formulation. ^22,23^ Because each denoising step produces an estimate of the clean mask, mask-level, structure-aware losses can be applied to it. The training loss is the same BCE + Dice used by all the trained baselines and is computed between the predicted mask 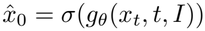 and the ground truth *x*_0_:

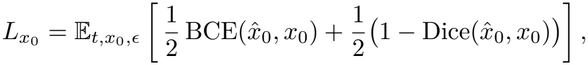

where *I* is the conditioning image and *σ* is the sigmoid.

### SAM

SAM was used as released, ^9^ with the ViT-B model (checkpoint sam_vit_b_01ec64), custom positive point prompting, and a mask threshold of 0.5. Unlike the other architectures, which take only the image, SAM also requires point prompts. To satisfy that, an intensity-based probability map was generated, and five points were chosen from high-probability regions on the basis of their number of neighboring points. SAM was run zero-shot with frozen weights, so no training occurred and only test metrics were computed. Prompting used positive points only, and all metrics were computed over the full resized image rather than within the bounding boxes, as done by Dr-SAM, ^12^ in order to maintain a fair comparison between all models.

### Convolutional and transformer baselines

All four of the following architectures were implemented with segmentation_models.pytorch, ^24^ initialized with random weights, and used a 3-channel input and 1-channel output with BCE+Dice loss over logits and the Adam optimizer.

ResNet-UNet uses a ResNet-50^25^ backbone, with an encoder and decoder each four convolutional blocks deep and skip connections ^26^ between corresponding stages to carry higher-resolution features, in a standard encoder →decoder forward pass.

ViT-DPT uses a ViT-B/16^18^ transformer encoder and a dense convolutional decoder following the DPT design. ^27^ In contrast to ResNet, tokens from several stages of the ViT are progressively combined and fed to the de-coder, so the decoder receives information from several resolutions directly.

DeepLabV3^28^ is built on a ResNet-50^25^ encoder with 8 × upsampling back to the input size. What makes it different is atrous convolution, where the receptive field is increased without any downsampling, along with atrous spatial pyramid pooling.

The hybrid ResNet-ViT architecture merges the local features from the ResNet ^25^ and the global feature context from the ViT. ^18^ It runs the ResNet-UNet and ViT-DPT networks described above in parallel on the same input, concatenates their output logits, and fuses them with a 1 × 1 convolution.

### Training protocol

All models were implemented in PyTorch, ^29^ with diffusers ^21^ or segmentation_models.pytorch ^24^ depending on the architecture, and trained on a single NVIDIA H100 GPU. All models except SAM were trained on all 25 seed-fold combinations at every data fraction with a , batch size of 16. At this batch size one epoch is 1, 2, or 5 optimizer updates at 16, 32, or 80 images, so each experiment included 50, 100, or 250 updates in total, equal for all models. Each ran for 50 epochs and was evaluated at the last epoch; no best-validation selection was applied and validation images were used only to track training. Table 1 documents the data fractions and the train/val/test sample counts they result in.

**Table 1.** Data fractions used for the data-efficiency study, with the resulting train/validation/test sample counts per run and the number of runs per model at each fraction (5 seeds, 5 folds each).

| **Data fraction** | **5%** | **10%** | **25%** |
|---|---|---|---|
| Train samples | 16 | 32 | 80 |
| Val samples | 4 | 8 | 20 |
| Test samples | 100 | 100 | 100 |
| Folds | 5 | 5 | 5 |
| Seeds | 5 | 5 | 5 |
| Runs per model | 25 | 25 | 25 |
| Test predictions | 2500 | 2500 | 2500 |

Mixed precision and gradient accumulation were not used. The Adam optimizer ^30^ was used with a learning rate of 10^*™*4^ and weight decay of 10^*™*5^. Apart from SAM, all encoders were initialized from random weights so that the comparison separates architecture from pretrained checkpoints.

### Evaluation and analysis

Images were randomly partitioned into 5 cross-validation folds, and this was repeated 5 times with random seeds; thus each (seed, fold) pair corresponds to a particular test fold of 100 images and a 400-image train+val pool to which the data fraction is applied. The same partitioning was used for all architectures and fractions. Test Dice and IoU scores were logged per image for each run, defined here as one experiment with a specific model, data fraction and (seed, fold) pair, meaning that each model-fraction combination generates 2,500 test predictions, in which every image occurs 5 times. Scores per run are averaged over the test fold; thus there are *n* = 25 pairs of runs per model and fraction.

For each metric and fraction we performed a one-sided paired *t*-test (two-sided 95% confidence interval on the mean difference, Cohen’s *d* ^31^), an exact sign-flip permutation test, and a one-sided Wilcoxon signed-rank test, ^32^ with rank-biserial correlation ^33^ as the effect size measure, on 25 pairs of *x*_0_-diffusion against each of the five competitors. The Wilcoxon *p*-values were Holm-corrected ^34^ across the five competitors within each metric and fraction, while a Friedman test ^35^ across all six models served as the omnibus test at each fraction. Two-sided Wilcoxon tests on the same pairs are reported alongside the one-sided results in Supplementary Table S1, and are the basis for statements of parity. Normality was verified with a Shapiro-Wilk test; ^36^ in case of failure, the Wilcoxon and sign-flip tests were considered the main result with parametric test results provided for comparison. The significance level was set to *α* = 0.05.

### Annotation timing

For estimating expert annotation time in hours and money, a vascular surgeon from our team labeled five angiograms from scratch without a model-predicted mask in Label Studio. ^37^ The five were test images from the first seed and fold of the 16-image *x*_0_-diffusion model. To get a better sense of the range of possible surgeondemand, the lowest, highest, and 3 average-scoring test images were used, evaluated using the test Dice against the ground truth, and because the five images were chosen and not sampled, the analysis is intended to be descriptive only.

The same surgeon also corrected the model’s predicted mask for each image, which was loaded into Label Studio as a pre-annotation. The surgeon labeling the images was already familiarized with Label Studio, and corrections were made on 24 October 2026 while from-scratch masks were drawn on a blank canvas on 3 November 2026, after an earlier from-scratch session on 12 September was stopped by a technical fault and its masks were discarded. More than seven weeks passed and unrelated masks were annotated between the technical fault and the final experiment, which should limit how much of each angiogram the surgeon remembered. The correction of the lowest-scoring image was repeated on 4 November while a fault in the Label Studio timer was investigated, and the repeat is the time reported.

For measuring time, we sourced the data from the lead times metric recorded by Label Studio. Finally, agreement was measured as the Dice between each corrected mask and the surgeon’s from-scratch mask of the same image. Campaign cost was projected as 16 from-scratch masks plus *N* − 16 corrected masks at the measured mean times. The labor cost was calculated using the 2024 average compensation of a vascular surgeon provided by Doximity, converted to an hourly rate on a 2,080-hour year. ^38^

## RESULTS

### Model performance by number of labels

Table 2 summarizes the six models at 16, 32, and 80 training images. A Friedman test over the six models rejected equality at each sample size (*p <* 10^−8^ for both metrics).

**Table 2.** Test Dice and IoU by number of labeled training images. Each cell is the mean (standard deviation) over the 25 runs (5 seeds, 5 folds) at that size. SAM is zero-shot and is scored on the same test folds without training.

|  | **Dice** |  |  | **IoU** |  |  |
|---|---|---|---|---|---|---|
| **Model** | **16 images** | **32 images** | **80 images** | **16 images** | **32 images** | **80 images** |
| *x*<sub>0</sub>-diffusion | 0.882 (0.017) | 0.883 (0.029) | 0.910 (0.032) | 0.795 (0.025) | 0.796 (0.043) | 0.840 (0.050) |
| SAM (zero-shot) | 0.835 (0.015) | 0.835 (0.015) | 0.835 (0.015) | 0.739 (0.018) | 0.739 (0.018) | 0.739 (0.018) |
| ViT-DPT | 0.791 (0.091) | 0.835 (0.072) | 0.908 (0.016) | 0.671 (0.108) | 0.729 (0.095) | 0.835 (0.025) |
| ResNet-ViT hybrid | 0.749 (0.161) | 0.830 (0.159) | 0.895 (0.096) | 0.630 (0.180) | 0.739 (0.181) | 0.823 (0.123) |
| ResNet-UNet | 0.703 (0.070) | 0.885 (0.016) | 0.923 (0.009) | 0.569 (0.074) | 0.802 (0.022) | 0.861 (0.013) |
| DeepLabV3 | 0.657 (0.103) | 0.895 (0.006) | 0.920 (0.005) | 0.518 (0.098) | 0.813 (0.008) | 0.854 (0.007) |

### Comparison of *x*_0_-diffusion with other models

Figure 2 shows the paired difference from *x*_0_-diffusion for each competitor. At 16 images every difference is positive and significant on both metrics, and *x*_0_-diffusion scored higher than each competitor in all 25 paired runs on Dice (*r* = +1.00, the smallest *p*-value possible after correction, 1.5 × 10^−7^). At 32 and 80 images only SAM and, at 32, ViT-DPT remain significantly behind; the small convolutional leads at 32 and 80 images are not significant in either direction (two-sided Wilcoxon, smallest Holm-adjusted *p* = 0.16, DeepLabV3 at 32 images on Dice; Supplementary Table S1). The paired differences failed the Shapiro-Wilk test in 25 of 30 comparisons, so Wilcoxon results are primary; the exact sign-flip and paired *t*-tests reached the same conclusion in every comparison (Supplementary Table S1).

**Figure 2.**
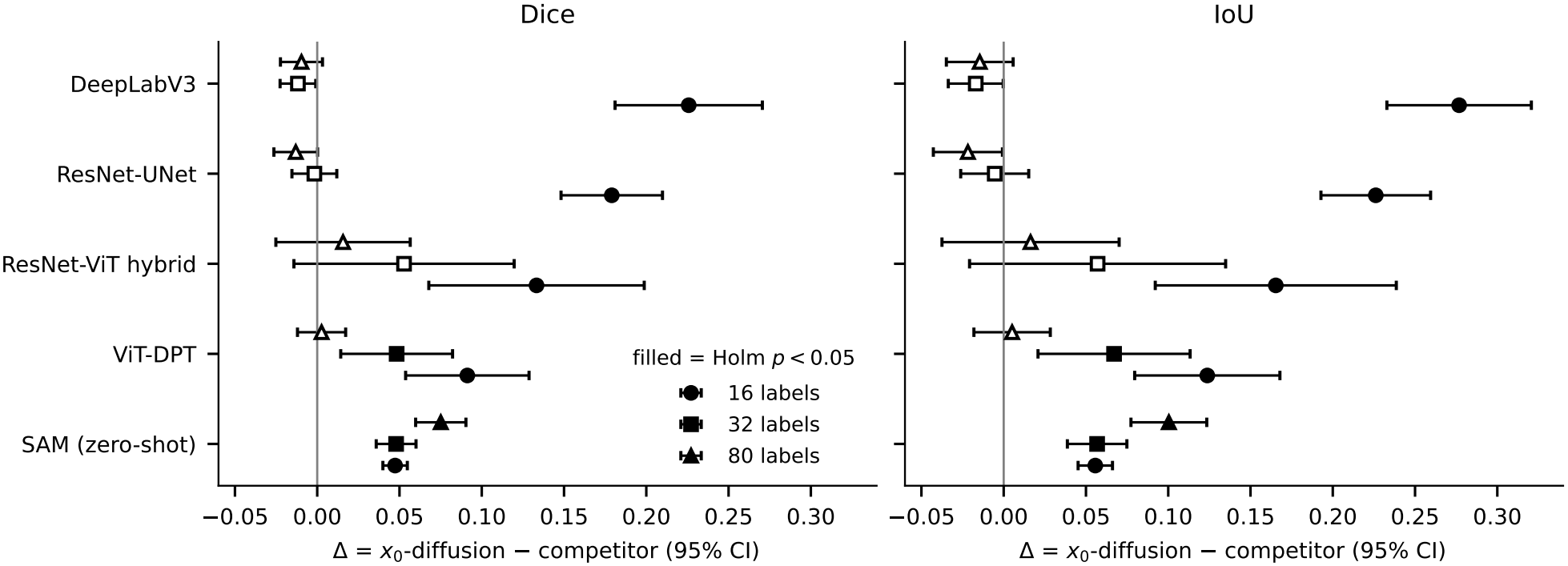
Paired difference in test Dice (left) and IoU (right) between *x*_0_-diffusion and each competitor at 16, 32, and 80 labeled training images. Points are the mean of (*x*_0_-diffusion minus competitor) over the 25 paired runs with 95% confidence intervals; values above zero favor *x*_0_-diffusion. Filled markers are significant on the one-sided Holm-corrected Wilcoxon test (*p <* 0.05). Full statistics, including rank-biserial correlations and two-sided tests, are in Supplementary Table S1. Alt text: Two forest plots. At 16 labels every competitor’s marker lies well to the right of zero with filled symbols; at 32 and 80 labels the ResNet-UNet and DeepLabV3 markers sit on the zero line with hollow symbols, while SAM stays to the right and filled at every size.

### Label efficiency

*x*_0_-diffusion achieved a Dice score of 0.882 with 16 training images, while the next two strongest baselines required twice as many labels to achieve this score. ResNet-UNet scored 0.885 and DeepLabV3 0.895 at 32 images, increasing from 0.703 and 0.657 at 16. Crossover occurs between 16 and 32 images and was not sampled. Beyond 32 images the convolutional models keep improving, reaching 0.92 Dice at 80 while *x*_0_-diffusion reaches 0.910 (Figure 3).

**Figure 3.**
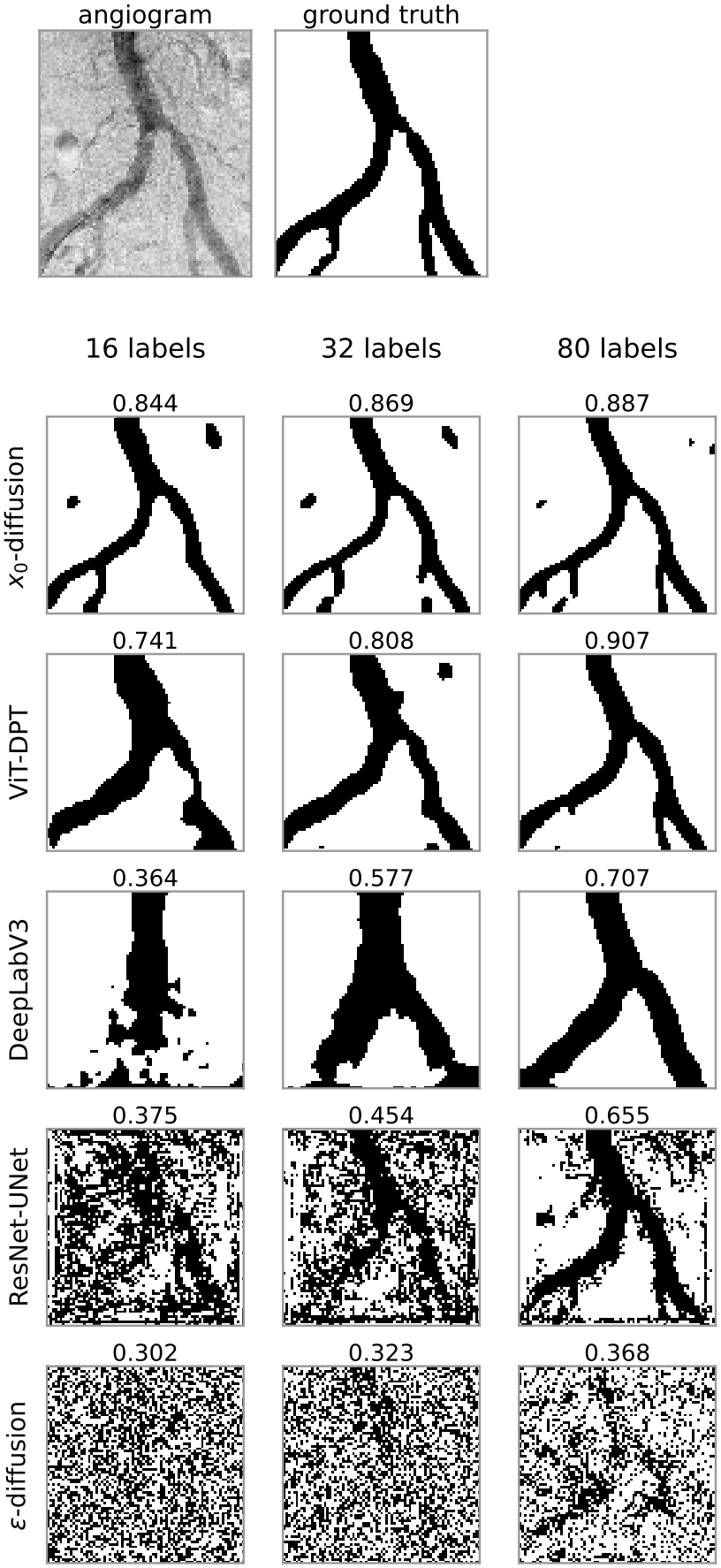
Predicted vessel masks for one test angiogram at 16, 32, and 80 labeled training images, with the source angiogram and expert mask above. Dice against the expert mask is shown over each panel. *x*_0_-diffusion recovers the vessel tree from 16 labels and changes little thereafter, whereas the convolutional baselines merge branches at 16 labels and improve steadily with more. The *ϵ*-prediction variant fails at every budget. Alt text: A grid of binary vessel masks. The *x*_0_-diffusion row shows a clean branching tree at all three label counts, closely matching the expert mask above; DeepLabV3 shows a merged blob at 16 labels that separates into branches by 80; the *ϵ*-diffusion row is speckled noise throughout.

### Prediction target ablation

The *ϵ*-prediction variant described in Methods scored 0.238, 0.232, and 0.230 Dice at 16, 32, and 80 training images, below every other model and with no improvement as labels were added (Supplementary Table S1). *x*_0_-diffusion beat it in all 25 paired runs at every fraction (rank-biserial +1.00; one-sided Wilcoxon *p* = 3 × 10^−8^, the smallest value 25 pairs can produce).

### Per-image behavior where zero-shot SAM fails

Run-level averages hide differences between models, so we compared per-image predictions of the 16-image models. SAM scored below 0.5 Dice on 136 predictions (5.4%). On the same images, *x*_0_-diffusion scored a median of 0.834 Dice (average 0.795) and ResNet-UNet an average of 0.571. *x*_0_-diffusion scored below 0.5 on only 8 predictions out of 2,500, and SAM failed on all 8. The tail of the distribution differs most (Figure 4): the 1st, 5th, and 10th percentiles of per-image Dice were 0.31, 0.49, and 0.63 for SAM and 0.70, 0.78, and 0.82 for *x*_0_-diffusion, and ResNet-UNet fell below 0.5 on 13.5% of predictions. Per-image scores were weakly correlated (Spearman *ρ* = 0.33), so a SAM failure does not predict an *x*_0_-diffusion failure, though the reverse holds, all 8 *x*_0_-diffusion failures were SAM failures.

**Figure 4.**
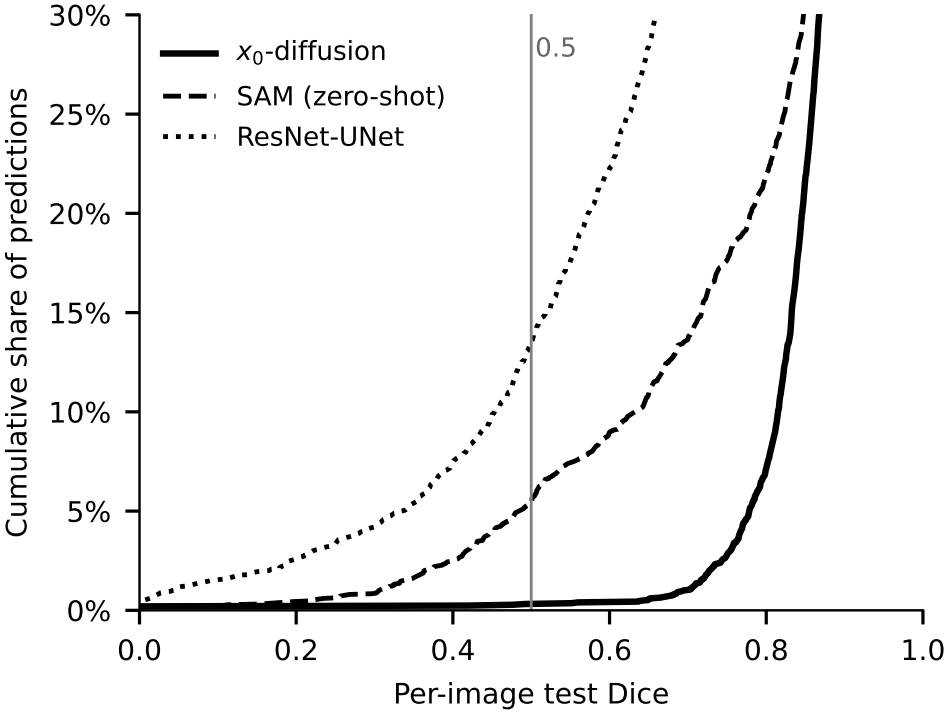
Cumulative distribution of per-image test Dice at 16 labeled training images over all 2,500 test predictions per model, restricted to the lower tail. Below the 0.5 threshold lie 0.3% of *x*_0_-diffusion predictions, 5.4% of zero-shot SAM, and 13.5% of ResNet-UNet. Alt text: A cumulative distribution plot. The *x*_0_-diffusion curve stays near zero until Dice 0.7, while the SAM and ResNet-UNet curves rise well before the 0.5 mark.

### Annotation time with and without model assistance

Correcting the model’s prediction was faster than annotating from scratch for all five images (Table 3). From-scratch annotation took a mean of 20.6 minutes per image (range 11.4 to 29.4) and correction took a mean of 11.1 minutes (range 7.4 to 17.1). The mean difference was 9.5 minutes (95% confidence interval −0.5 to 19.6), a 46% reduction in mean time. The saving did not follow prediction quality: it was 59% for the lowest-Dice prediction and 15% for the highest. Corrected masks agreed with the surgeon’s own from-scratch masks at Dice 0.963 to 0.984 (mean 0.972).

**Table 3.**
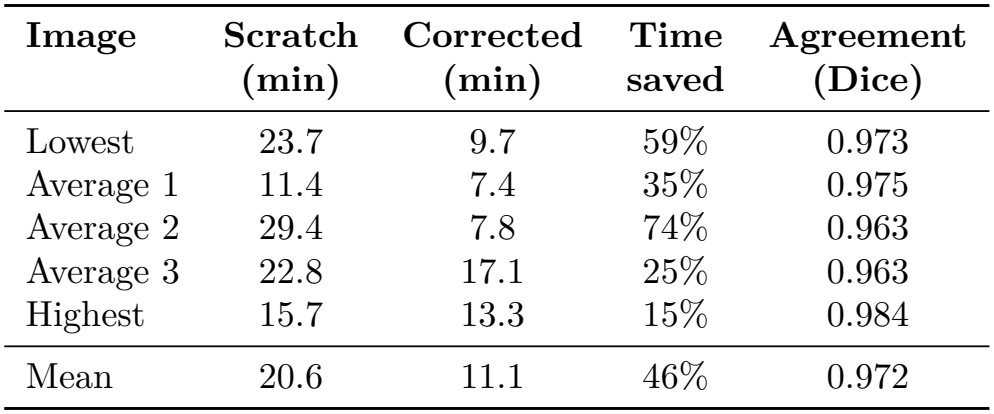
Time for one vascular surgeon to annotate five test angiograms from scratch and by correcting the prediction of the 16-image *x*_0_-diffusion model. Lowest, average, and highest refer to the prediction’s test Dice against the Dr-SAM reference. Agreement is the Dice between the corrected and from-scratch masks. Time saved is the per-image reduction; the last row gives the reduction in mean time and the mean agreement.

## DISCUSSION

### *x*_0_-diffusion’s advantage under label sparsity

At 16 training images, *x*_0_-diffusion outperformed all other models on both metrics, though the advantage was confined to that regime (Table 2 and Figure 2). We propose three hypotheses, of which only the third is partly tested here (the prediction target ablation).

Firstly, corrupting the mask for 1000 timesteps provides the network with exposure to a wide range of reconstruction problems, while predicting the clean mask directly means that they become tasks of reconstructing vascular structures rather than predicting denoising patterns. This matters most at 16 training images, where the other networks see too little variety to learn from and the corruption procedure works as a form of implicit regularization. Secondly, iterative denoising of the mask with conditioning on the image at each step gives the model many opportunities to correct the mistakes made locally, as opposed to a single forward pass. Thirdly, the *x*_0_ parameterization allows for the use of structure-aware loss functions, such as the Dice + BCE combined loss employed, making the training signal more geometrically consistent with the mask itself. The diffusion UNet also has more parameters than either convolutional baseline (Methods), so lower capacity does not explain its advantage at 16 images.

### Ablation of the prediction target

The *ϵ*-prediction variant performed far worse than every other model at every fraction (Results). We attribute this to the MSE loss rather than to the *ϵ* parameterization itself. A noise objective will result in a uniform loss gradient across the whole image and will not create any sharp boundary line between the vessels and the background. SegDiff and MedSegDiff train with *ϵ*-prediction successfully, ^16,17^ and so under a larger training budget the gap would likely narrow; under our matched budget, *x*_0_ prediction is what allows Dice and BCE to be applied, and we recommend it for diffusion-based segmentation of fine vascular structure.

### SAM performs poorly even with scale

Although SAM ^9^ was trained on one billion masks and is often used zero-shot, *x*_0_-diffusion achieves superior results at each fraction probed, and the gap widens as *x*_0_-diffusion sees more labeled data (Figure 2). We hypothesize that this occurs due to a domain shift, where the training data used for SAM (SA-1B) comprises mostly natural objects containing abundant texture and distinct edges, which do not match the thin and branching low-contrast appearance of angiographic vessels. ^39^ A secondary factor may be the prompting strategy. Points are selected based on intensity differences, and while this is effective for clean angiograms, this breaks down when there is noise or distortion present in the angiogram, leading to degenerate masks.

Our zero-shot SAM reaches 0.739 mean IoU, below the 0.859 reported by the Dr-SAM authors for their full positive and negative point selection pipeline. ^12^ The two are not directly comparable, as they compute IoU within the expert-annotated bounding boxes on 450 images, while we compute it over the full image on all 500, and our re-implementation prompts with positive points only.

### Starting a labeling campaign

Our findings suggest that *x*_0_-diffusion could anchor a label-efficient segmentation strategy such as would be needed for a labeling campaign. Trained on a small seed set, it gives annotators predicted masks to correct, and it can be retrained as corrected masks accumulate, the standard active-learning loop. ^40,41^ Failures are rare enough for this to be feasible, only 8 of 2,500 predictions from the 16-image model fell below 0.5 Dice.

The cost of a labeling campaign can be estimated in surgeon-hours and dollars. In our timing study, one mask took the surgeon a mean of 20.6 minutes from scratch and 11.1 minutes by correction (Table 3). The 2024 average vascular surgery compensation is $576,452, ^38^ which is about $277 per hour, and thus one from-scratch mask costs around $95 of surgeon time and one corrected mask around $51. Annotating the 80 images at which the convolutional baselines reach 0.92 Dice would take 27.5 hours ($7,600) from scratch, and annotating all 500 Dr-SAM images would require 172 hours ($47,600) (Table 4). Annotating 16 seed images from scratch and correcting the rest would instead take 17.3 hours ($4,800) and 94.7 hours ($26,200), which is 37% and 45% less surgeon time.

**Table 4.** Projected vascular surgeon time and cost to annotate 80 or 500 angiograms, either all from scratch or with 16 seed images from scratch and the rest by correcting *x*_0_-diffusion predictions. Projections use the measured means of 20.6 minutes per from-scratch mask and 11.1 minutes per corrected mask (Table 3) and $277 per hour.

|  | **From scratch** |  | *x*<sub>0</sub>**-assisted** |  |  |
|---|---|---|---|---|---|
| **Labels** | Hours | Cost | Hours | Cost | **Saving** |
| 80 | 27.5 | \$7,600 | 17.3 | \$4,800 | 37% |
| 500 | 171.7 | \$47,600 | 94.7 | \$26,200 | 45% |

These figures come from five images and a single surgeon, and thus are a first estimate (see Limitations). Corrected masks matched the surgeon’s own from-scratch masks at a mean Dice of 0.97, so correction did not lower quality by the annotator’s own standard. The projection also holds the 16-image model fixed. Retraining on corrected masks could shorten later corrections, which we did not test.

### Limitations

Several issues limit the conclusions of this study. The first is the dataset, Dr-SAM covers one modality, one anatomical region (the infrarenal aorta and iliac arteries), and two centers, and its distribution is narrow, so the data efficiency of *x*_0_-diffusion may not transfer to other vessels, regions, or modalities, including the belowknee arteries where CbA has the most diagnostic value. Most Dr-SAM images are natively 386 × 448, and the 50 that are 819 × 950 scored no worse after downsampling (mean Dice within 0.014 of the smaller images for every model at every training-set size), so the 384 × 384 input forced by ViT compatibility cost little here, though other resolutions could shift the trends.

Second, the sample, the *n* = 25 runs in each cell reuse the same 500 images under different partitions and are not independent, and the primary tests are one-sided in favor of *x*_0_-diffusion. As noted in Results, a two-sided test finds no significant difference at 32 or 80 images in either direction (Supplementary Table S1).

Since both the epochs and batch size remained constant, the number of optimizer updates scaled with respect to the number of labels available. Within each training-set size the budget is identical across architectures, so the comparisons in Table 2 and Figure 2 are budget-matched and the 16-image result is not con-founded. However, the trend across sizes is confounded. When the baselines improve from 16 to 32 images they receive both twice the labels and twice the updates, so our data cannot separate label efficiency from tolerance of a short optimization budget. Training longer at 16 images would raise the risk of overfitting, and that risk differs by architecture, ^42^ so the two factors cannot be disentangled without an equal-update experiment at every size, which we did not run.

The timing study is a pilot, and as stated before, it used five images chosen to span prediction quality and one surgeon. Both conditions used the same images. Four corrections were made 10 days before the from-scratch masks, and all five images had been seen in the discarded first session, so familiarity favored the from-scratch condition and the measured saving is probably conservative. For the same reason the two masks of each image are not independent, and their agreement may partly reflect anchoring to the model’s mask. They were compared only with each other, not with the Dr-SAM reference. The correction time for the lowest-scoring image comes from a repeat, and lead times include inspection and any idle time. The projection assumes that the mean times hold across a campaign.

The comparison set was chosen to isolate inductive bias under matched conditions rather than to benchmark the strongest available system for this task, and three omissions follow from that. All encoders except SAM were randomly initialized, so ImageNet or medical pretrained convolutional baselines, which a laboratory would use in practice, may close the 16-image gap. We did not use any data augmentation, which may have adversely affected the performance of discriminative models more than a diffusion model, which implicitly involves perturbing the mask. Dedicated diffusion-based segmentation frameworks like SegDiff, MedSegDiff, ^16,17^ and even foundational models like MedSAM ^10^ finetuned or prompted for a particular task were not implemented in this work. Comparing to them, along with pretraining and augmentation, is the next natural study.

## CONCLUSION

The data show that *x*_0_-diffusion is the best performing architecture for label efficiency in angiogram segmentation, and this holds across all 25 paired experiments on Dice. Its lead fades as more labels are introduced, so this, together with its resistance to the catastrophic failures of zero-shot SAM, means that its best use case is as a bootstrapping step in an aggressive labeling campaign. In a five-image pilot, correcting its predictions took a vascular surgeon 46% less time than annotating from scratch, which projects to 37% to 45% fewer surgeon-hours over campaigns of 80 to 500 labels.

## Data Availability

This study used ONLY openly available human data that were originally located at https://github.com/vazgenzohranyan/Dr.SAM. No new patient data were collected. The annotation timing involved only members of the study team, so institutional review board approval was not required.

https://github.com/vazgenzohranyan/Dr.SAM/tree/main

## ETHICS STATEMENT

This study used the publicly available, de-identified Dr-SAM dataset ^12^ and generated no new patient data. Annotation timing involved only members of the study team, and no human subjects research was conducted, so institutional review board approval was not required.

## DATA AVAILABILITY

The Dr-SAM dataset is a publicly available dataset. ^43^ Code for all experiments, along with other project information, is available at https://github.com/ALCHEMI-Labs/Label-efficient-vessel-segmentation

## AUTHOR CONTRIBUTIONS

Neelay Ranjan: Conceptualization, Methodology, Software, Formal analysis, Investigation, Data curation, Visualization, Writing – original draft, Writing – review and editing. Shantanu Dev: Methodology, Software, Data curation, Formal analysis, Writing – review and editing, Supervision. Andrew Alexander Gonzalez: Conceptualization, Investigation (expert annotation), Resources, Writing – review and editing, Supervision.

## ACKNOWLEDGMENTS

The authors used Claude (Anthropic) for code development, for reviewing the reported statistics against the raw run data, and for suggestions on language. All content was explicitly vetted and approved by the authors.

## SUPPLEMENTARY MATERIAL

Supplementary Table S1 is provided as a supplementary file. It gives the paired statistics for all 30 comparisons and for the *ϵ*-diffusion ablation, including the exact signflip and paired *t*-test results and the post hoc two-sided Wilcoxon test.

