## Supplementary Table 1 for "Bootstrapping surgeon labeling campaigns with x_0_-diffusion: label-efficient vessel segmentation of catheter-based angiograms"

**Supplementary Table S1.** Paired comparison of every competitor with $x_0$-diffusion on the $n = 25$ paired runs per cell (5 seeds × 5 folds). $\Delta$ is the mean of ($x_0$-diffusion minus competitor) with its 95% confidence interval; positive values favor $x_0$-diffusion. $d$ is Cohen's $d$ for paired samples. One-sided tests ($t$-test, exact sign-flip permutation test, and exact Wilcoxon signed-rank test) test the hypothesis that $x_0$-diffusion is superior; $r$ is the rank-biserial correlation of the Wilcoxon test. Holm correction is applied across the five competitors within each metric and training-set size. The post hoc two-sided Wilcoxon test referenced in the Limitations is given in the last two columns. At $\alpha = 0.05$, the one-sided $t$-test, sign-flip test, and Holm-corrected Wilcoxon test reach the same conclusion in all 30 comparisons. $\epsilon$-diffusion, the prediction-target ablation, is compared with the same paired tests but is not part of the Holm family, so its Holm columns are marked with a dash; its mean (SD) over the 25 runs at 16, 32, and 80 images is 0.238 (0.033), 0.232 (0.041), 0.230 (0.037) Dice and 0.136 (0.021), 0.132 (0.027), 0.131 (0.024) IoU.

| | | | One-sided tests | | | | | Two-sided Wilcoxon | |
|---|---|---|---|---|---|---|---|---|---|
| **Competitor** | $\Delta$ [95% CI] | $d$ | $p_t$ | $p_{\text{sign}}$ | $p_W$ | $p_W$ (Holm) | $r$ | $p_W$ | $p_W$ (Holm) |
| **Dice, 16 labeled training images** | | | | | | | | | |
| SAM (zero-shot) | 0.047 [0.040, 0.055] | 2.63 | $9.0 \times 10^{-13}$ | $3.0 \times 10^{-8}$ | $3.0 \times 10^{-8}$ | $1.5 \times 10^{-7}$ | 1.00 | $6.0 \times 10^{-8}$ | $3.0 \times 10^{-7}$ |
| ViT-DPT | 0.091 [0.054, 0.129] | 1.00 | $2.0 \times 10^{-5}$ | $3.0 \times 10^{-8}$ | $3.0 \times 10^{-8}$ | $1.5 \times 10^{-7}$ | 1.00 | $6.0 \times 10^{-8}$ | $3.0 \times 10^{-7}$ |
| ResNet-ViT hybrid | 0.133 [0.068, 0.199] | 0.84 | $1.6 \times 10^{-4}$ | $3.0 \times 10^{-8}$ | $3.0 \times 10^{-8}$ | $1.5 \times 10^{-7}$ | 1.00 | $6.0 \times 10^{-8}$ | $3.0 \times 10^{-7}$ |
| ResNet-UNet | 0.179 [0.148, 0.210] | 2.40 | $6.4 \times 10^{-12}$ | $3.0 \times 10^{-8}$ | $3.0 \times 10^{-8}$ | $1.5 \times 10^{-7}$ | 1.00 | $6.0 \times 10^{-8}$ | $3.0 \times 10^{-7}$ |
| DeepLabV3 | 0.226 [0.181, 0.271] | 2.08 | $1.1 \times 10^{-10}$ | $3.0 \times 10^{-8}$ | $3.0 \times 10^{-8}$ | $1.5 \times 10^{-7}$ | 1.00 | $6.0 \times 10^{-8}$ | $3.0 \times 10^{-7}$ |
| $\epsilon$-diffusion (ablation) | 0.644 [0.630, 0.659] | 18.31 | $2.4 \times 10^{-32}$ | $3.0 \times 10^{-8}$ | $3.0 \times 10^{-8}$ | – | 1.00 | $6.0 \times 10^{-8}$ | – |
| **Dice, 32 labeled training images** | | | | | | | | | |
| SAM (zero-shot) | 0.048 [0.036, 0.060] | 1.64 | $1.1 \times 10^{-8}$ | $8.0 \times 10^{-7}$ | $1.6 \times 10^{-6}$ | $8.2 \times 10^{-6}$ | 0.93 | $3.3 \times 10^{-6}$ | $1.6 \times 10^{-5}$ |
| ViT-DPT | 0.048 [0.014, 0.082] | 0.59 | 0.004 | 0.002 | 0.001 | 0.006 | 0.66 | 0.003 | 0.011 |
| ResNet-ViT hybrid | 0.053 [−0.014, 0.120] | 0.33 | 0.058 | 0.068 | 0.644 | 1.000 | −0.08 | 0.731 | 1.000 |
| ResNet-UNet | −0.002 [−0.015, 0.012] | −0.05 | 0.602 | 0.597 | 0.396 | 1.000 | 0.06 | 0.791 | 1.000 |
| DeepLabV3 | −0.012 [−0.023, −0.001] | −0.45 | 0.982 | 0.987 | 0.976 | 1.000 | −0.45 | 0.052 | 0.155 |
| $\epsilon$-diffusion (ablation) | 0.651 [0.632, 0.671] | 13.86 | $1.8 \times 10^{-29}$ | $3.0 \times 10^{-8}$ | $3.0 \times 10^{-8}$ | – | 1.00 | $6.0 \times 10^{-8}$ | – |
| **Dice, 80 labeled training images** | | | | | | | | | |
| SAM (zero-shot) | 0.075 [0.060, 0.090] | 2.03 | $1.8 \times 10^{-10}$ | $2.1 \times 10^{-7}$ | $2.1 \times 10^{-7}$ | $1.0 \times 10^{-6}$ | 0.98 | $4.2 \times 10^{-7}$ | $2.1 \times 10^{-6}$ |
| ViT-DPT | 0.003 [−0.012, 0.017] | 0.07 | 0.356 | 0.364 | 0.132 | 0.527 | 0.26 | 0.263 | 0.790 |
| ResNet-ViT hybrid | 0.016 [−0.025, 0.057] | 0.16 | 0.218 | 0.256 | 0.644 | 1.000 | −0.08 | 0.731 | 0.884 |
| ResNet-UNet | −0.013 [−0.026, 0.000] | −0.41 | 0.973 | 0.979 | 0.940 | 1.000 | −0.35 | 0.127 | 0.507 |
| DeepLabV3 | −0.010 [−0.022, 0.003] | −0.31 | 0.932 | 0.933 | 0.787 | 1.000 | −0.18 | 0.442 | 0.884 |
| $\epsilon$-diffusion (ablation) | 0.681 [0.663, 0.698] | 15.87 | $7.2 \times 10^{-31}$ | $3.0 \times 10^{-8}$ | $3.0 \times 10^{-8}$ | – | 1.00 | $6.0 \times 10^{-8}$ | – |
| **IoU, 16 labeled training images** | | | | | | | | | |
| SAM (zero-shot) | 0.056 [0.045, 0.066] | 2.20 | $3.7 \times 10^{-11}$ | $6.0 \times 10^{-8}$ | $6.0 \times 10^{-8}$ | $1.5 \times 10^{-7}$ | 0.99 | $1.2 \times 10^{-7}$ | $3.0 \times 10^{-7}$ |
| ViT-DPT | 0.124 [0.080, 0.168] | 1.16 | $2.9 \times 10^{-6}$ | $3.0 \times 10^{-8}$ | $3.0 \times 10^{-8}$ | $1.5 \times 10^{-7}$ | 1.00 | $6.0 \times 10^{-8}$ | $3.0 \times 10^{-7}$ |
| ResNet-ViT hybrid | 0.165 [0.092, 0.239] | 0.93 | $4.9 \times 10^{-5}$ | $3.0 \times 10^{-8}$ | $3.0 \times 10^{-8}$ | $1.5 \times 10^{-7}$ | 1.00 | $6.0 \times 10^{-8}$ | $3.0 \times 10^{-7}$ |
| ResNet-UNet | 0.226 [0.193, 0.259] | 2.80 | $2.4 \times 10^{-13}$ | $3.0 \times 10^{-8}$ | $3.0 \times 10^{-8}$ | $1.5 \times 10^{-7}$ | 1.00 | $6.0 \times 10^{-8}$ | $3.0 \times 10^{-7}$ |
| DeepLabV3 | 0.277 [0.233, 0.321] | 2.60 | $1.2 \times 10^{-12}$ | $3.0 \times 10^{-8}$ | $3.0 \times 10^{-8}$ | $1.5 \times 10^{-7}$ | 1.00 | $6.0 \times 10^{-8}$ | $3.0 \times 10^{-7}$ |
| $\epsilon$-diffusion (ablation) | 0.659 [0.646, 0.671] | 21.52 | $4.9 \times 10^{-34}$ | $3.0 \times 10^{-8}$ | $3.0 \times 10^{-8}$ | – | 1.00 | $6.0 \times 10^{-8}$ | – |
| **IoU, 32 labeled training images** | | | | | | | | | |
| SAM (zero-shot) | 0.057 [0.039, 0.075] | 1.30 | $5.3 \times 10^{-7}$ | $5.8 \times 10^{-6}$ | $7.5 \times 10^{-6}$ | $3.8 \times 10^{-5}$ | 0.89 | $1.5 \times 10^{-5}$ | $7.5 \times 10^{-5}$ |
| ViT-DPT | 0.067 [0.021, 0.113] | 0.60 | 0.003 | 0.002 | 0.002 | 0.006 | 0.66 | 0.003 | 0.012 |
| ResNet-ViT hybrid | 0.057 [−0.021, 0.135] | 0.30 | 0.072 | 0.077 | 0.683 | 1.000 | −0.11 | 0.653 | 1.000 |
| ResNet-UNet | −0.005 [−0.026, 0.015] | −0.11 | 0.705 | 0.698 | 0.584 | 1.000 | −0.05 | 0.853 | 1.000 |
| DeepLabV3 | −0.017 [−0.034, −0.000] | −0.42 | 0.977 | 0.980 | 0.965 | 1.000 | −0.41 | 0.075 | 0.226 |
| $\epsilon$-diffusion (ablation) | 0.664 [0.644, 0.683] | 13.93 | $1.6 \times 10^{-29}$ | $3.0 \times 10^{-8}$ | $3.0 \times 10^{-8}$ | – | 1.00 | $6.0 \times 10^{-8}$ | – |
| **IoU, 80 labeled training images** | | | | | | | | | |
| SAM (zero-shot) | 0.100 [0.077, 0.123] | 1.80 | $1.9 \times 10^{-9}$ | $3.9 \times 10^{-7}$ | $3.0 \times 10^{-7}$ | $1.5 \times 10^{-6}$ | 0.97 | $6.0 \times 10^{-7}$ | $3.0 \times 10^{-6}$ |
| ViT-DPT | 0.005 [−0.018, 0.028] | 0.09 | 0.327 | 0.333 | 0.132 | 0.527 | 0.26 | 0.263 | 0.790 |
| ResNet-ViT hybrid | 0.016 [−0.038, 0.070] | 0.13 | 0.269 | 0.289 | 0.654 | 1.000 | −0.09 | 0.711 | 0.852 |
| ResNet-UNet | −0.022 [−0.043, −0.001] | −0.43 | 0.979 | 0.983 | 0.957 | 1.000 | −0.39 | 0.090 | 0.361 |
| DeepLabV3 | −0.015 [−0.035, 0.006] | −0.30 | 0.924 | 0.922 | 0.795 | 1.000 | −0.19 | 0.426 | 0.852 |
| $\epsilon$-diffusion (ablation) | 0.709 [0.688, 0.730] | 14.06 | $1.3 \times 10^{-29}$ | $3.0 \times 10^{-8}$ | $3.0 \times 10^{-8}$ | – | 1.00 | $6.0 \times 10^{-8}$ | – |
